# Exploring clinical care pathways of individuals with comorbid mental health disorders after presenting to emergency due to acute musculoskeletal pain: A Narrative Review

**DOI:** 10.1101/2024.10.18.24315727

**Authors:** Priya Arora, James Michael Elliott, Fereshteh Pourkazemi

## Abstract

Complex Musculoskeletal (MSK) pain conditions are the leading cause of Years Lived with Disability (YLD) globally [1]. Alarmingly, this has remained the same since 1990 [2] suggesting that research into prevention and rehabilitation of MSK pain over the past 25+ years has had limited effect on its overall global burden. The reasons some fail to report full recovery while others follow a less problematic recovery trajectory are becoming clearer with psychological predictors (anxiety, depression, stress) showing some prognostic value [3]. Effective interventions however have proven elusive. Treatment of chronic MSK pain in many clinical settings tends to focus on the physical modalities such as pharmacologic, surgical, and other physical therapies excluding holistic interventions targeting psychosocial causes [4]. An integrative approach towards assessing and effectively managing a patient’s pain should cover the physical, behavioural, and psychosocial drivers of the patient’s pain experience. Moreover, a better understanding of the myriad of biopsychosocial mechanisms driving the clinical course for each patient seems particularly germane to the acute care encounter, given the current challenges with pharmaceutical dependence and overutilisation of and reliance on diagnostic tests that rarely inform management; or worse, promote ineffective management.

## INTRODUCTION

Nine out of ten individuals will report and seek care for their pain following an acute injury involving the musculoskeletal (MSK) system. Recovery from acute MSK injury is expected for the majority, but the same cannot be expected, nor observed, for all, with nearly 2-3 out of 10 transitioning from acute to chronic pain [5]. Individuals’ recovery pathways are undoubtedly influenced by factors that extend well beyond the original pain producing injury. In fact, a simple question arises when two people exposed to the same external perturbation (e.g., another car hitting their car while stopped at a red light), have different pathways towards recovery. One may recover spontaneously, while the other may take longer…or may never offer words to suggest they feel, and believe they are, recovered. What factors drive this prolonged experience, and why? Appreciating the rhetorical nature of these questions, one can (and should) consider the many factors influencing these two very different recovery pathways (and disparate therapeutic approaches on offer) following the ‘same’ injury. [6].

To date, current clinical pathways for acute musculoskeletal (MSK) injury have existed in silos. These pathways typically manage the most common MSK traumatic injuries, namely those to the cervical or lumbar spine and lower limbs, presenting to emergency departments (ED). The current protocols focus on triage to exclude any major distracting injury (e.g., head injury, fracture, spinal cord injury). Once traumatic distracting injury has been excluded or triaged, the focus shifts to a diagnosable injury with management options aimed to discharge with an appropriate medication recommendation, follow up advice or discharge support to primary care or allied health services. These discharge options can influence the clinical course, and in some, not all, cases, increase risk for adverse effects.[7]

Individual guidelines [8, 9] exist for MSK trauma in primary care, but these are not targeted to prognostic risk in the primary care, post-discharge, environment. The disparate experience in the primary care encountered is often driven by professional scope-of-practice and guidelines that exist for each body regions and condition…and for each profession [10]. For example, across Australia, appropriate/consistent advice on management of non-traumatic low back pain is provided in only 20% of GP consultations [11]. In neck pain disorders, over 50% of people receive, what is considered, unnecessary imaging [12], and substantial gaps exist between evidence-based guidelines and clinical management of lower limb injuries[13]. It has been suggested that nearly all care pathways fail to consider prognostic risk [14], resulting in delayed care for those at higher risk (poorer prognosis) and care for those at lower risk (good prognosis), considered to be ineffective, and in some cases, harmful [15, 16].

This non-systematic literature review aims to examine the evidence on care pathways of individuals presenting to an emergency department due to acute MSK pain, with pre-existing comorbid mental health (M.H.) disorders and for a reported mental illness (M.I.).

## METHODS

### Database search

The authors undertook a comprehensive search across six electronic databases of MEDLINE, PubMed, PsycINFO, Embase, CINAHL and Google Scholar, using the keywords and search strategy, provided in table 1. The search strategy was modified where required, for other databases.

**Table 1:**
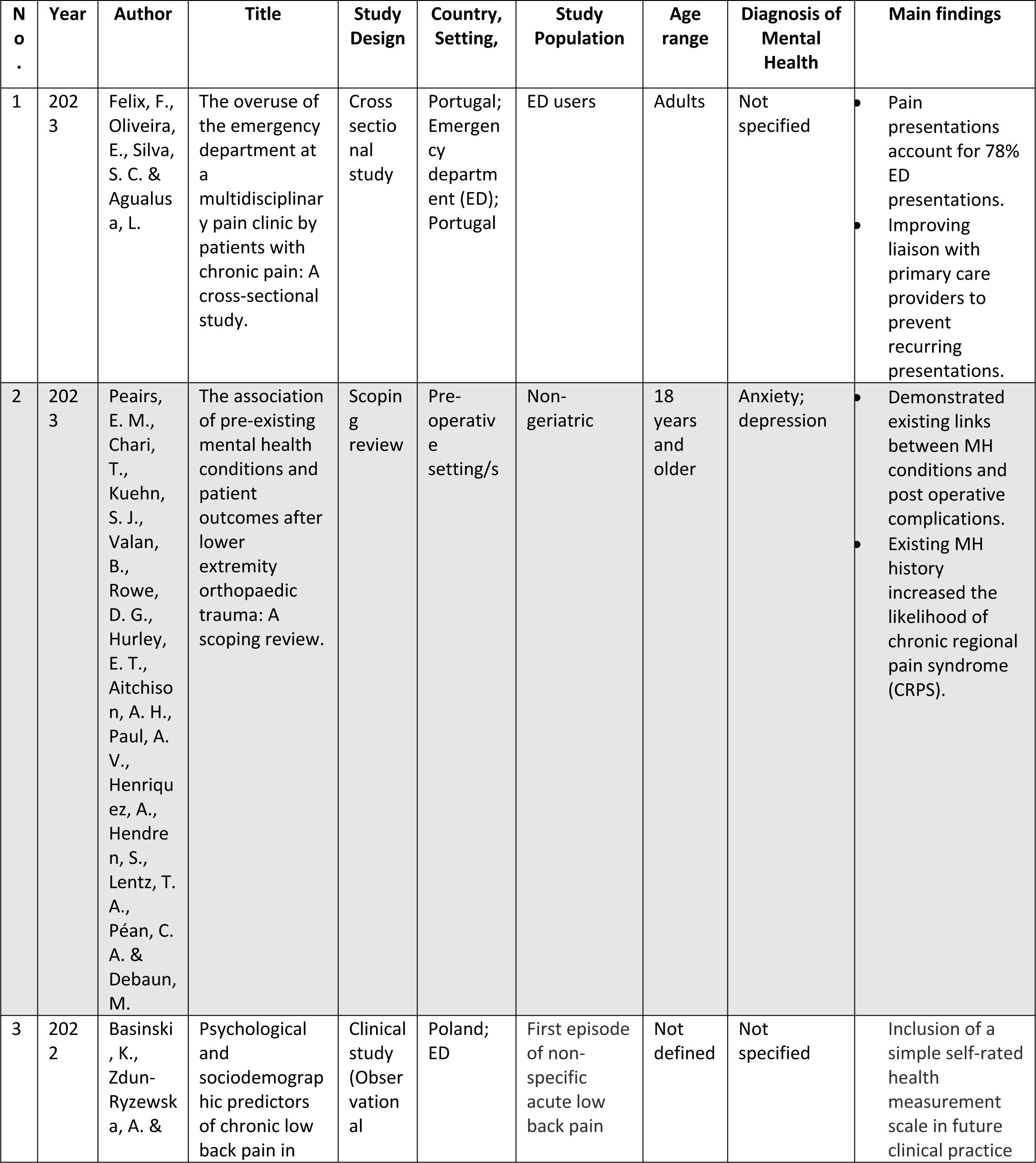
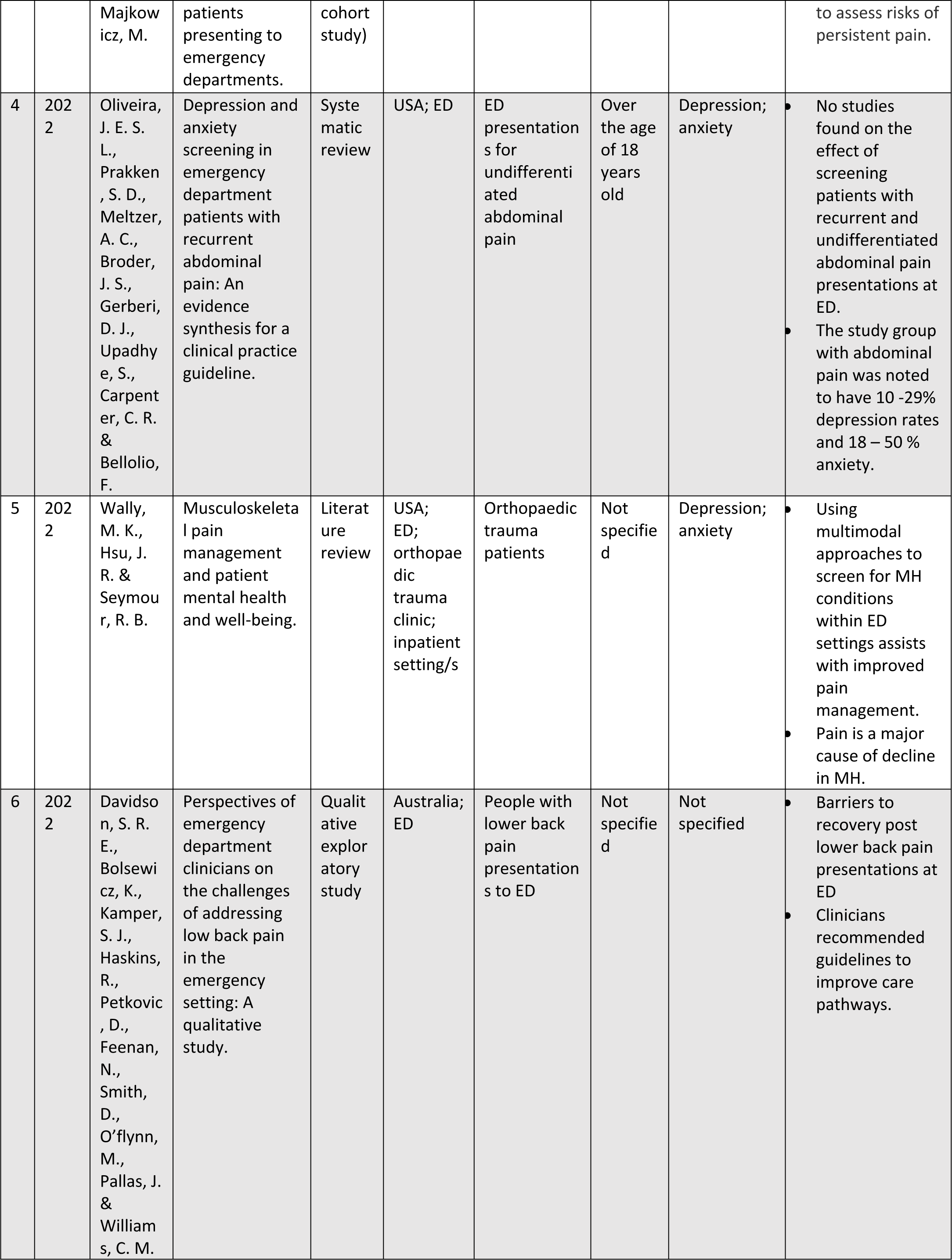
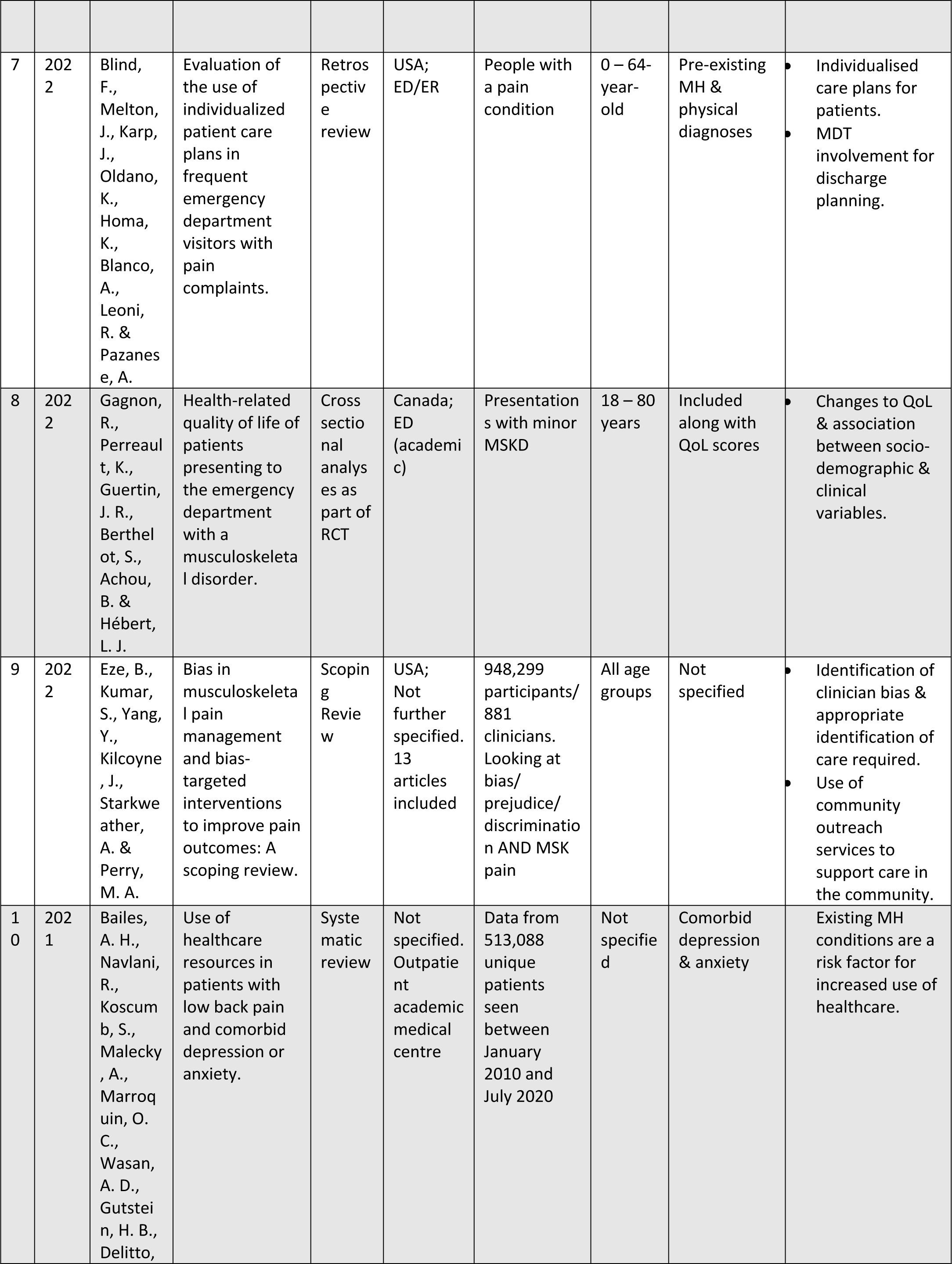
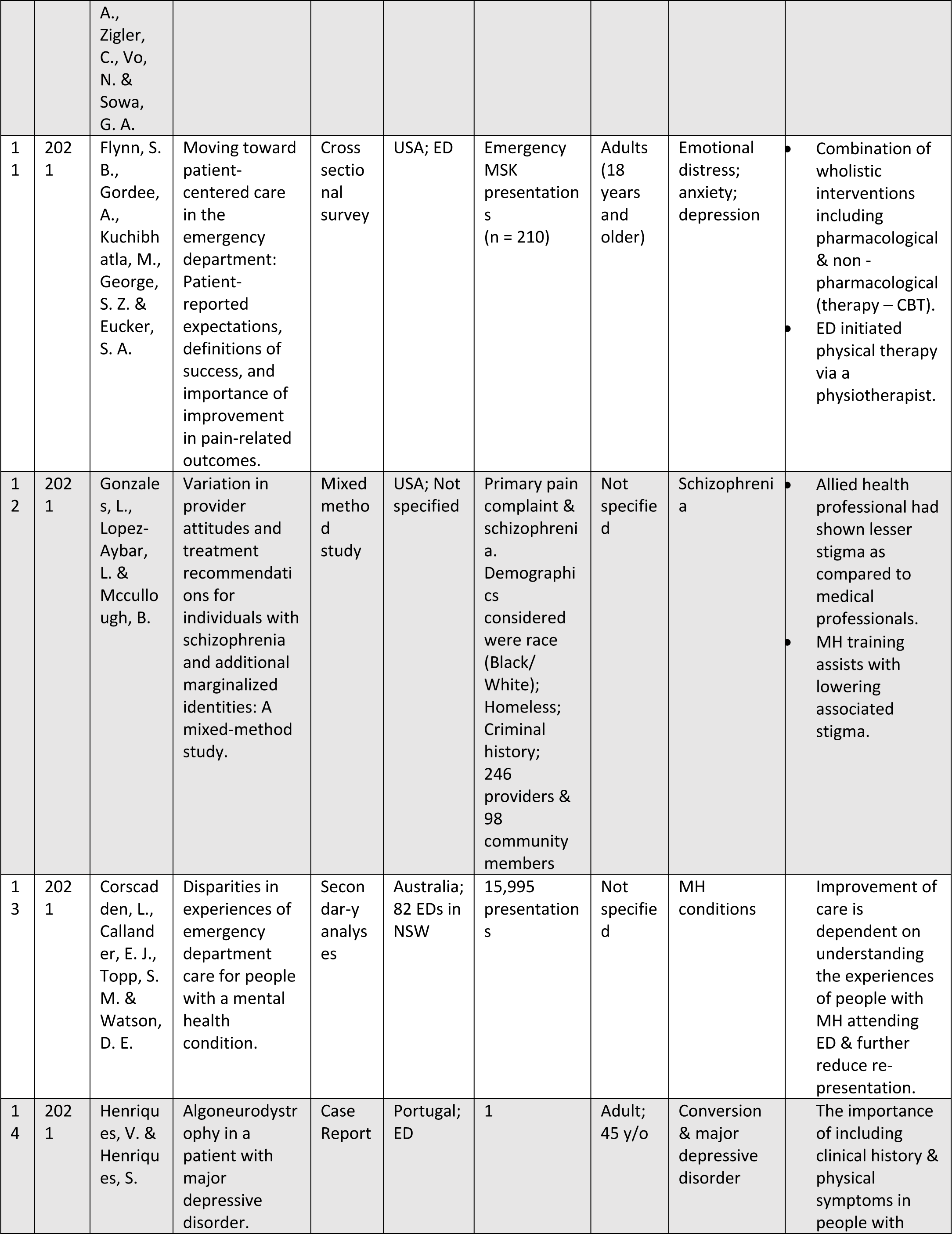
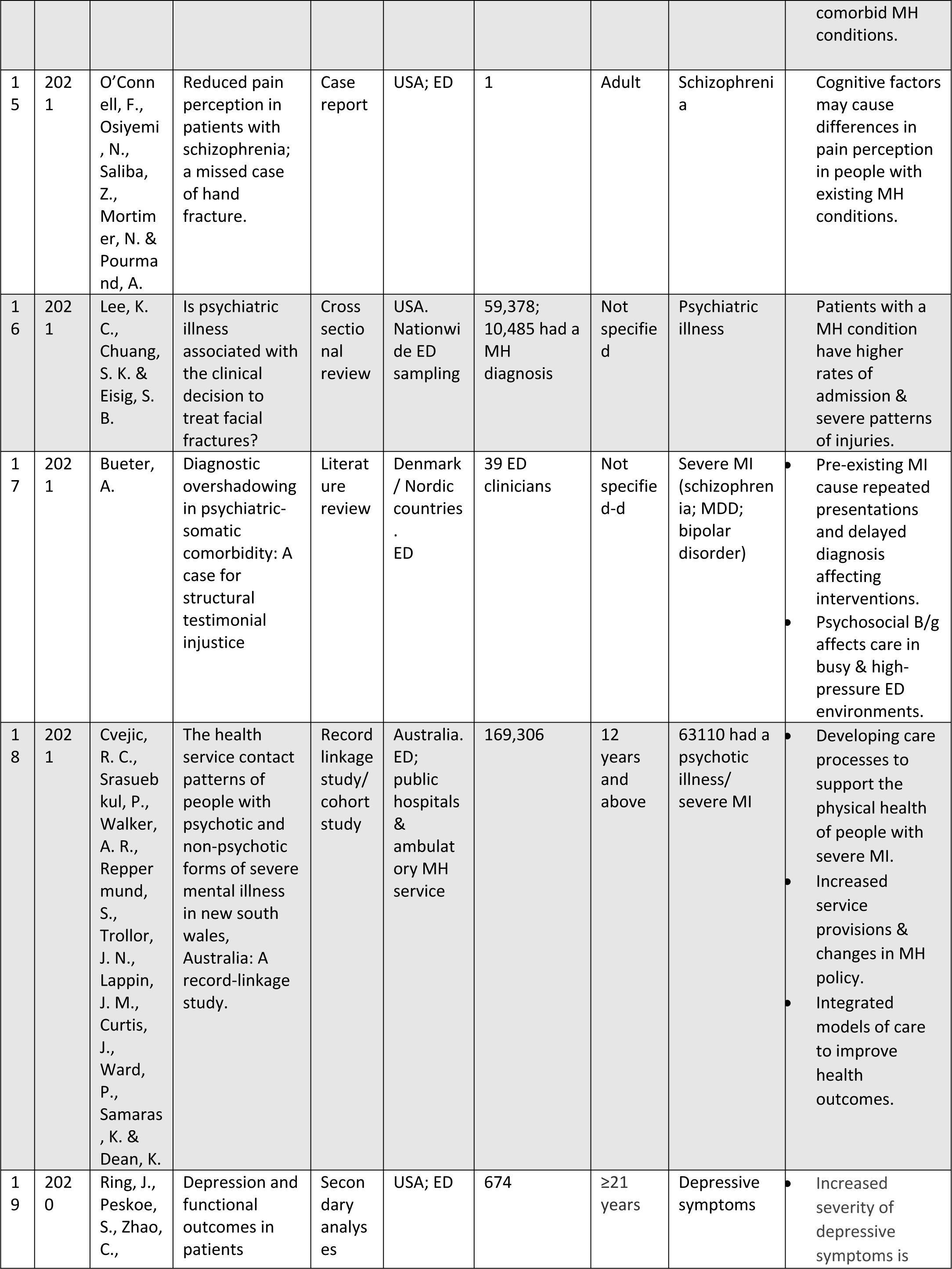
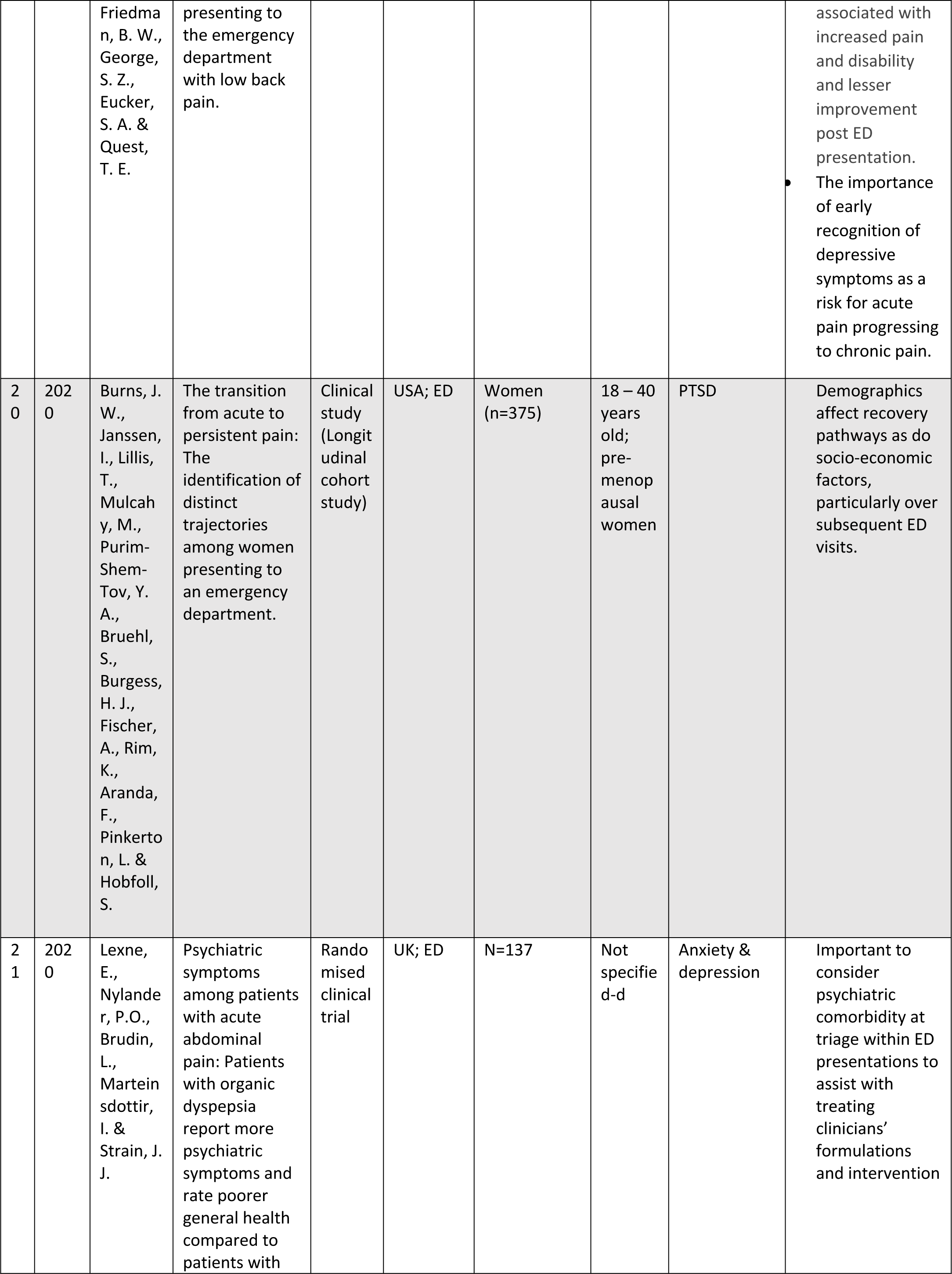
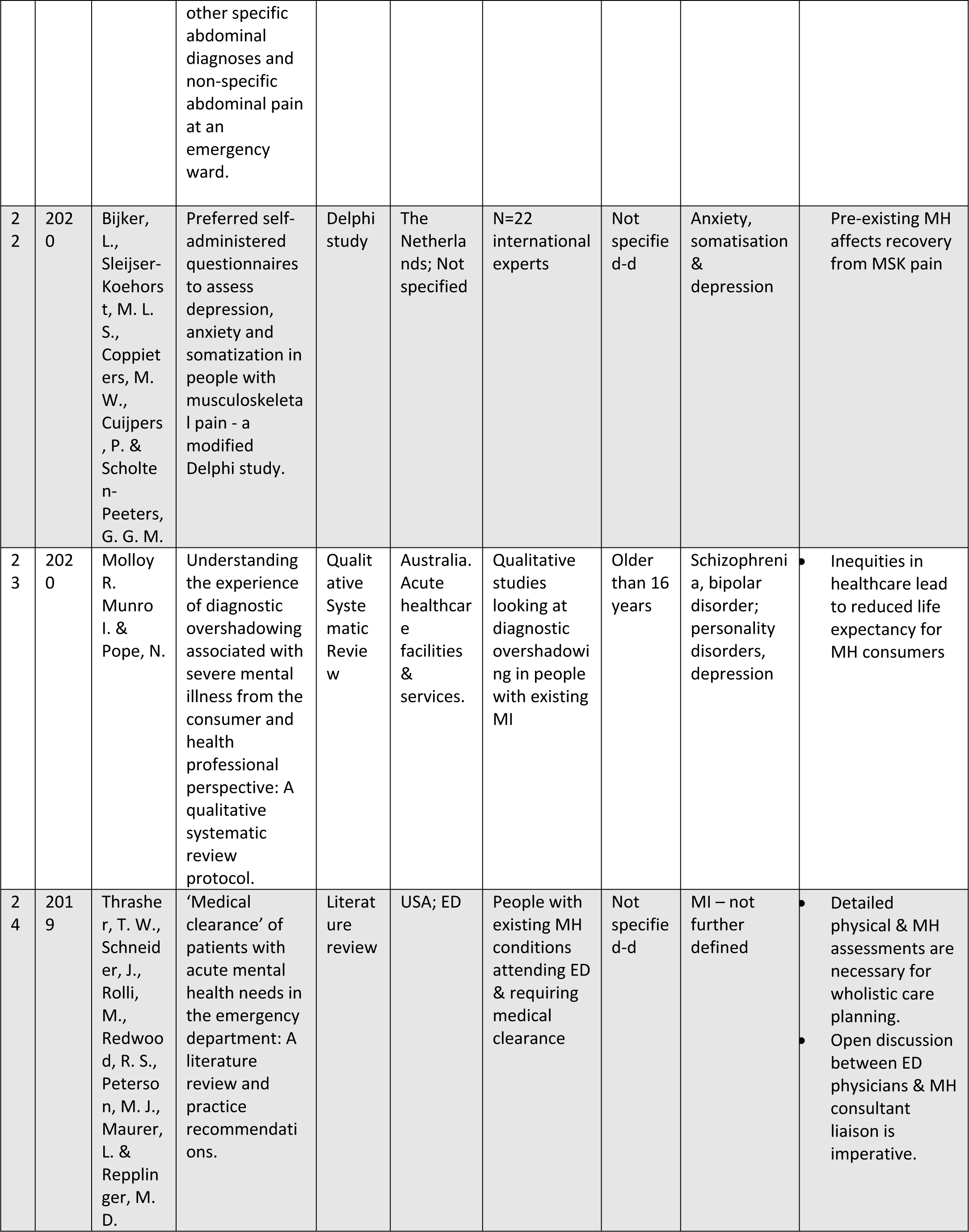
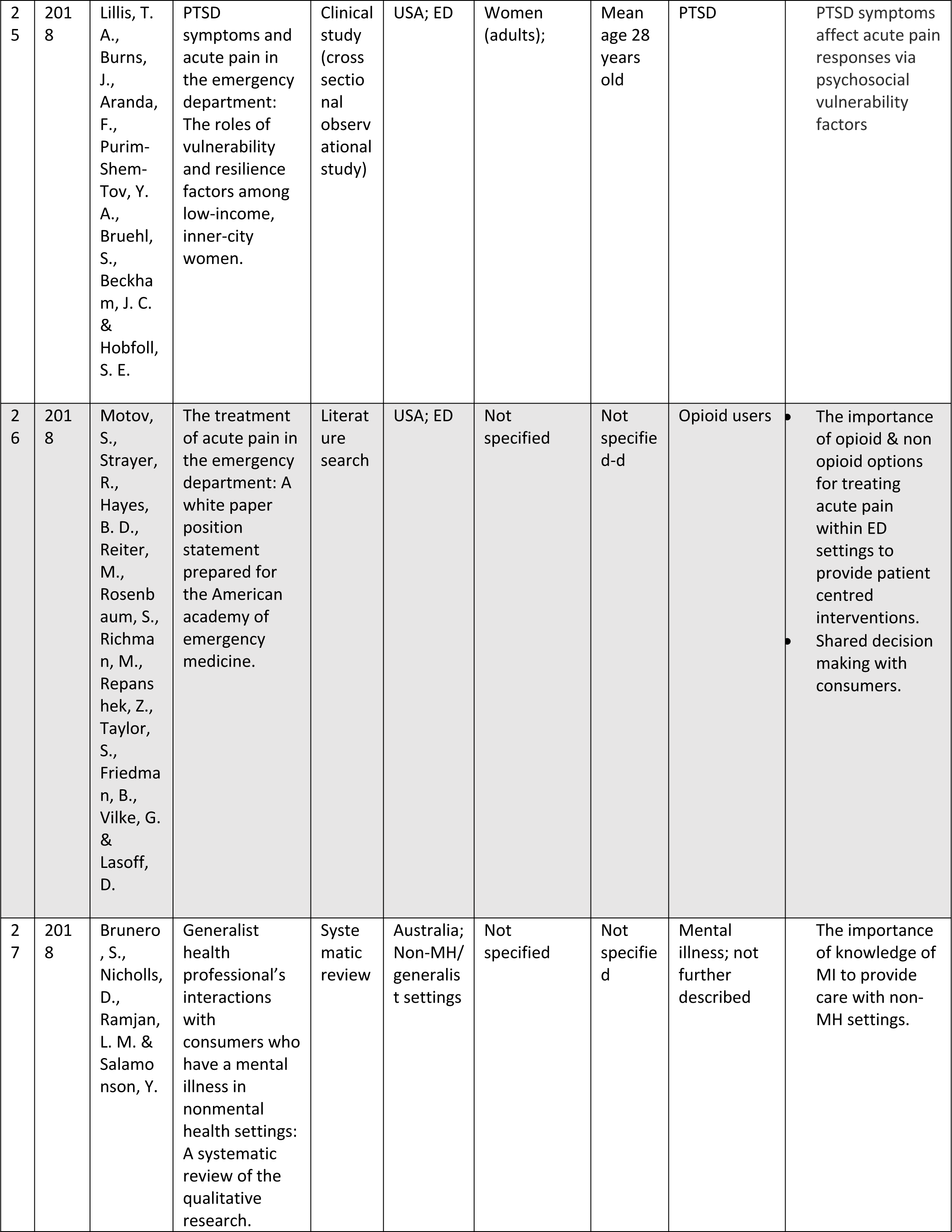
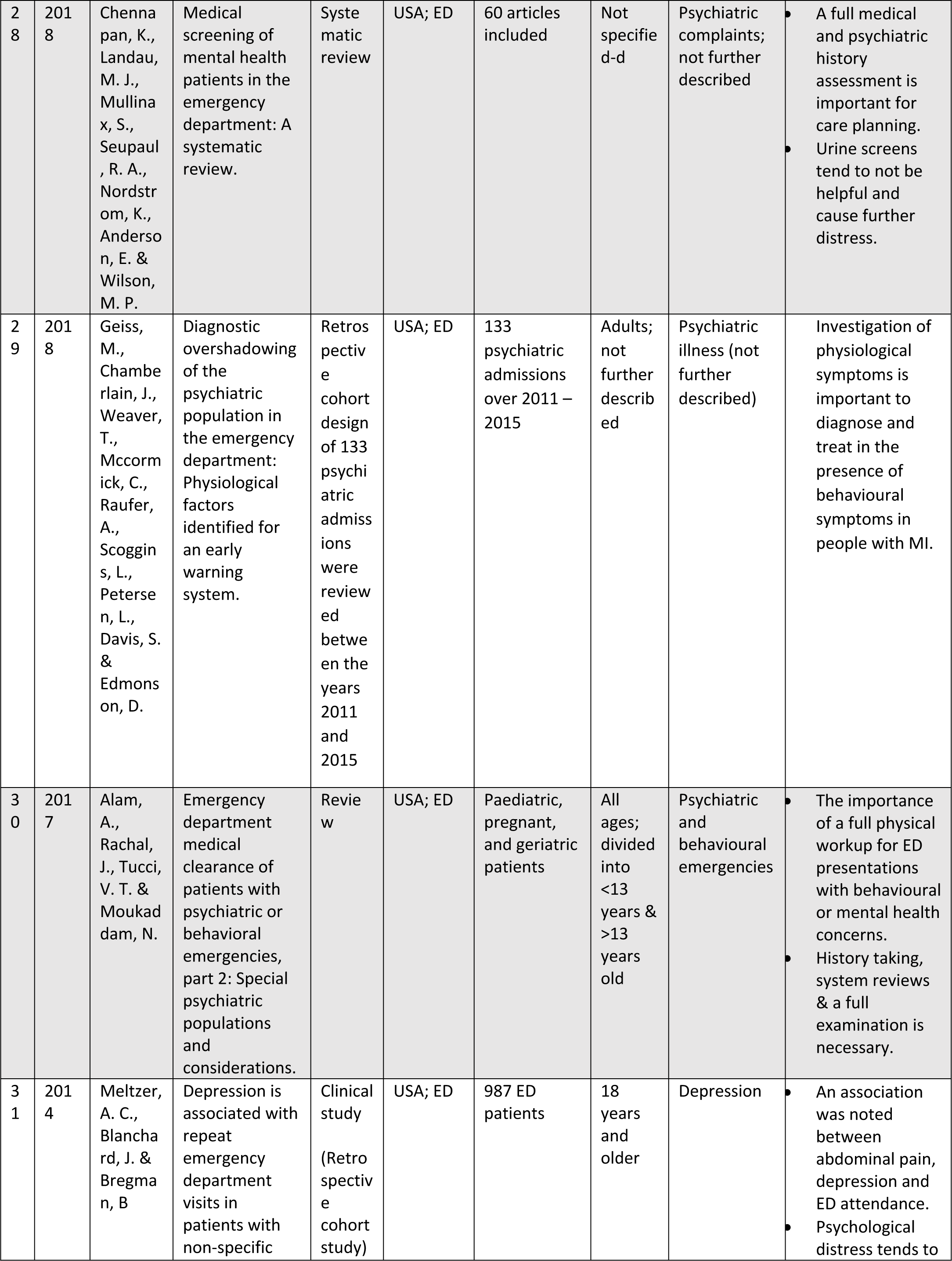
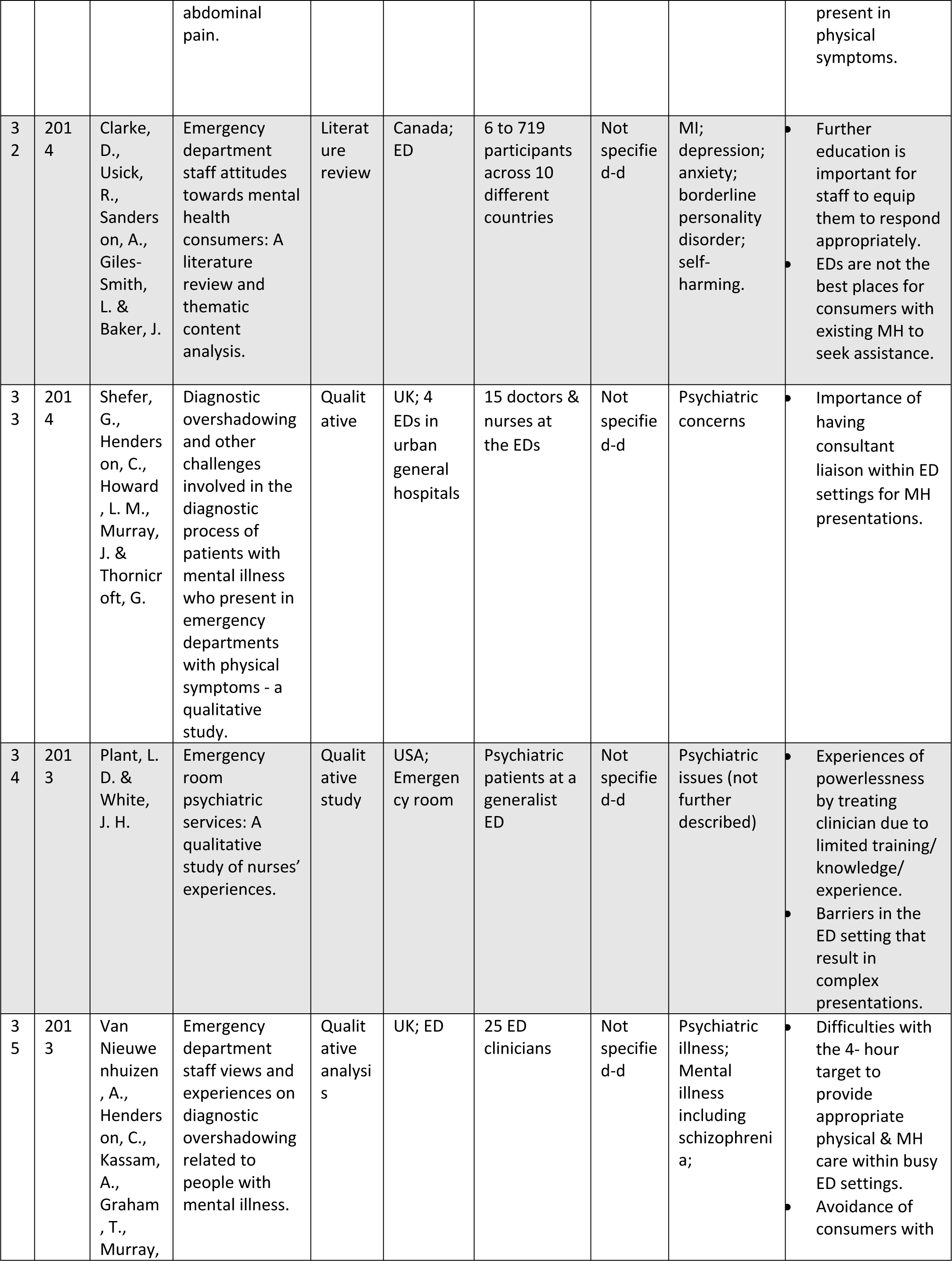
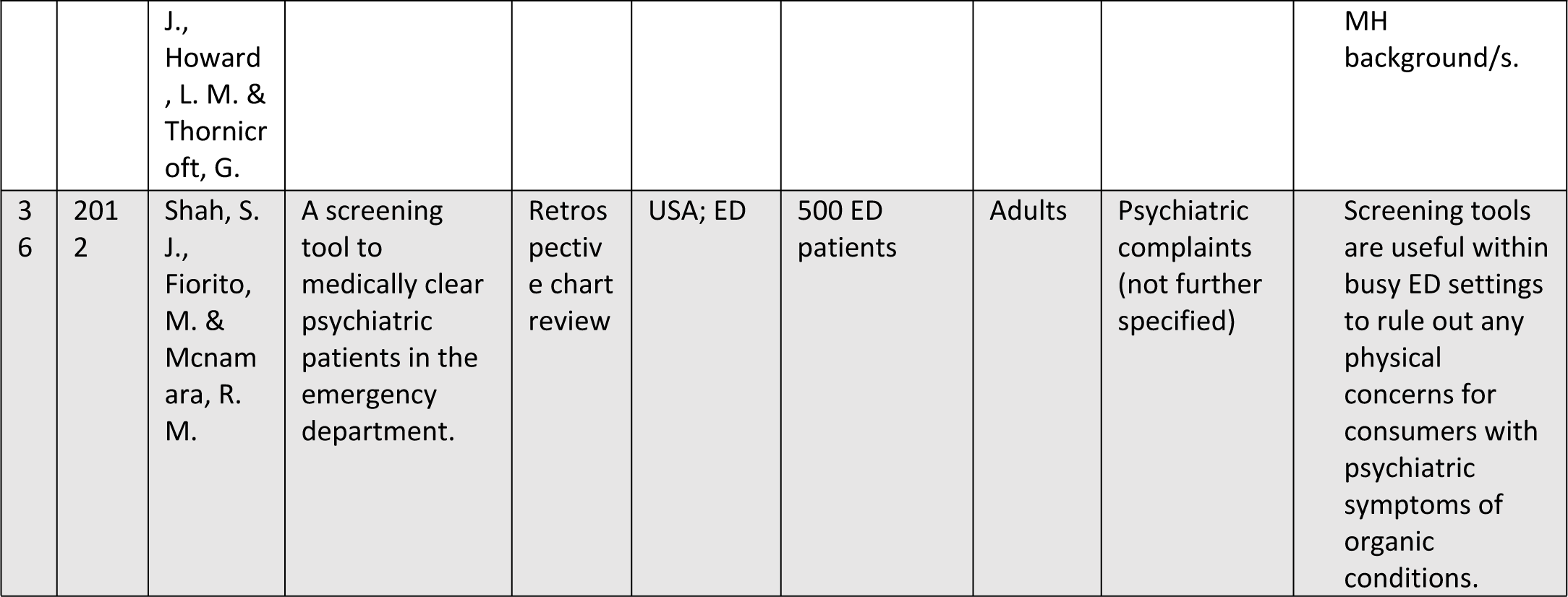
Overview of Literature Pertaining to Emergency Department Visits for Acute Pain Complaints in People with Mental Health Conditions.

Inclusion criteria: studies were included if they were full published peer-reviewed studies written in English since 2010 onwards. The participants were adults (18 years and older) with acute MSK pain presentations. The study setting was at the emergency department, looking at the clinical pathways for the people presenting with mental health issues as compared to others.

Exclusion criteria: studies were excluded if the publications captured non-MSK presentations; drug & alcohol related issues; traumatic injuries; cognitive impairment; requiring surgical interventions; not clearly defined age limits.

In consultation with an expert librarian, a literature search was completed using the following terms. The search strategy was modified accordingly, in each database to include MSK pain, systems and disorders along with mental illness or disorder with emergency settings/ hospitals/ services looking at adult populations from 2010 onwards. At the conclusion of the search, results were exported to EndNote citation software for screening.

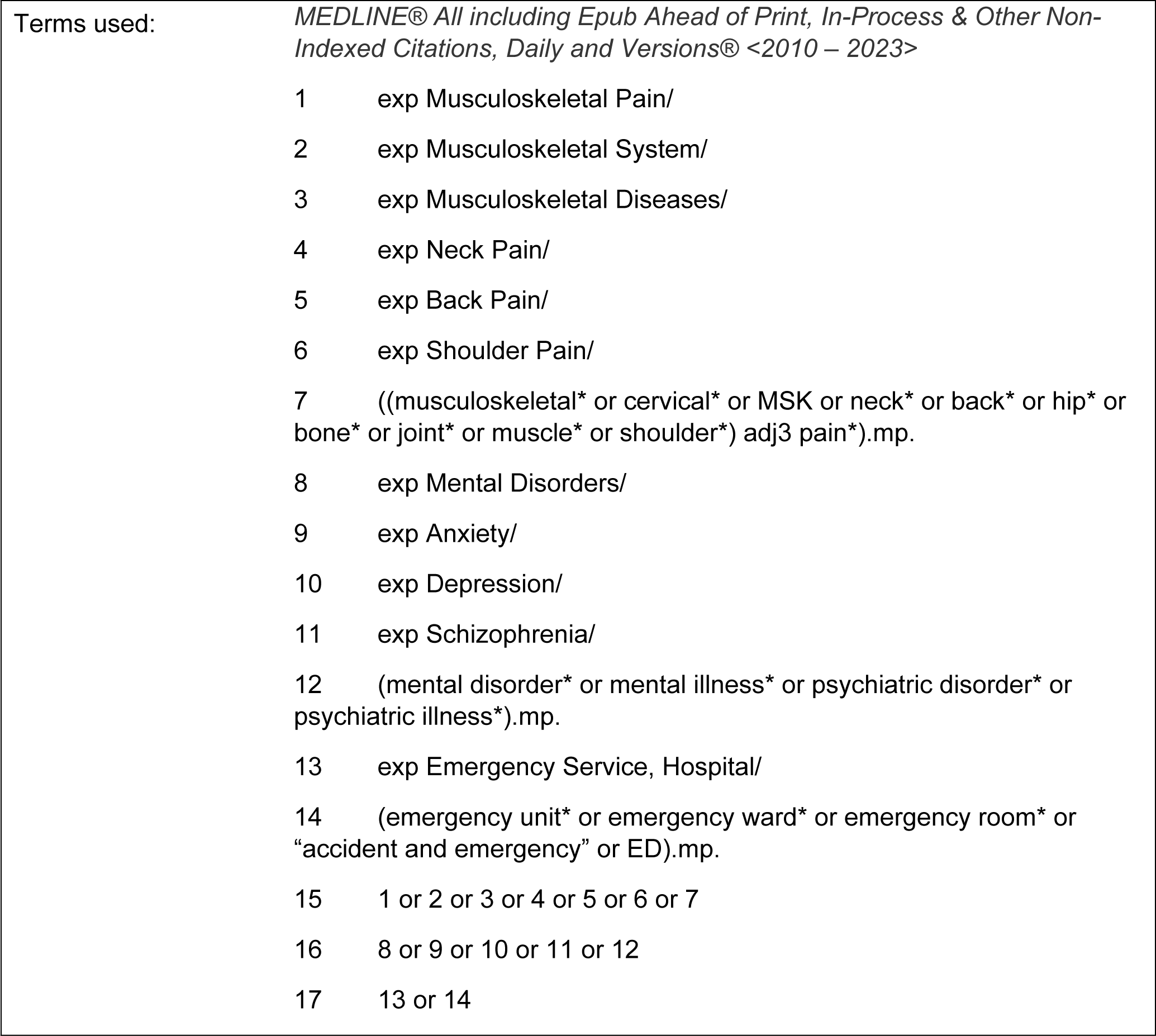
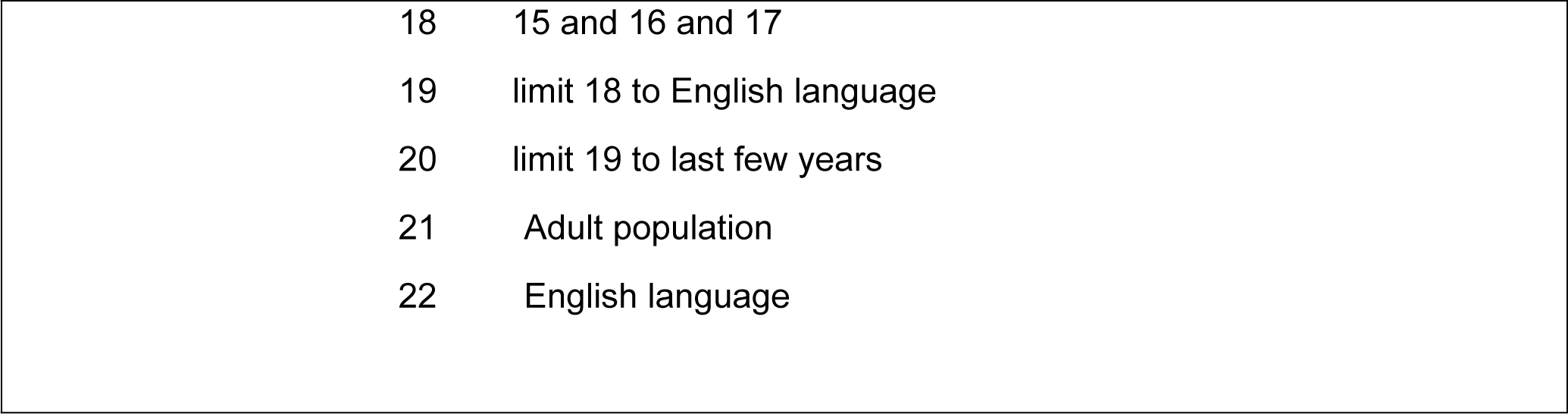

### Literature Screening

The first author (PA) screened studies based on the inclusion criteria, by going through titles and abstracts, and then full papers. The following data items were extracted from the included studies comprising of study characteristics including the year, author, setting, participants demographics, intervention and care pathways (where reported). Identified publications and articles were synthesised using narrative synthesis. The narrative synthesis was conducted using the structure provided by Popay, et al [17]. Exploring themes of findings involves identifying common patterns and insights across studies on interventions in acute settings.

Our search strategy identified 75 papers and after screening against inclusion criteria, 36 studies were included for data extraction and reporting in our narrative review (Figure 1).

**Figure 1:**
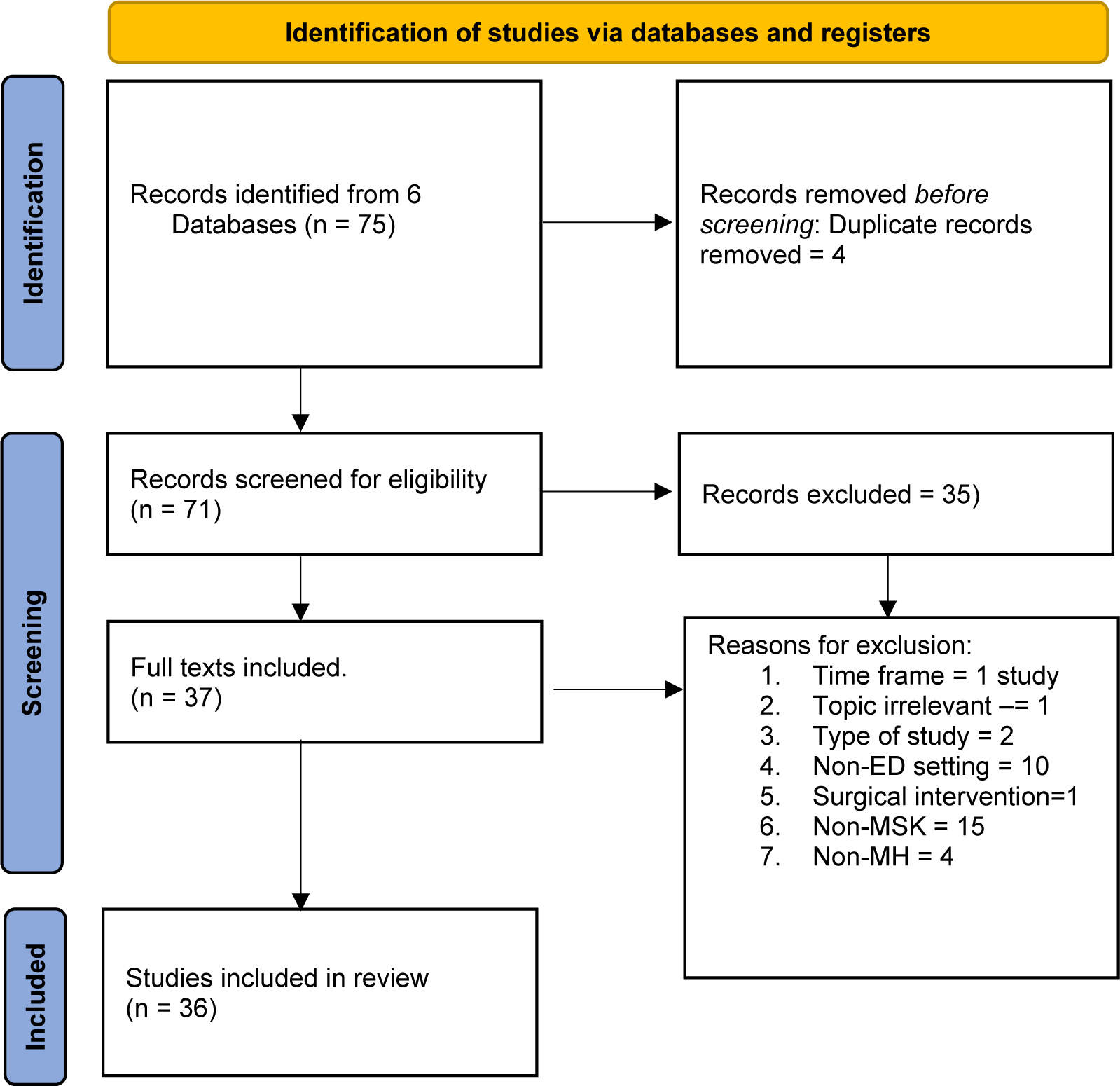
Flow diagram of studies.

The included studies were a combination of systematic (n=6), literature (n=4) or scoping (n=2) reviews, cross-sectional qualitative (n=5) or quantitative (n=4) studies, retrospective chart reviews (n=3), case studies/reports (n=3), clinical linkage studies (n=1), clinical studies (n=4), a randomised clinical study, and a Delphi study. Further two studies included secondary analysis of data from survey-based studies; one across 82 EDs in New South Wales (NSW)), and another from approximately 15,000 participants rating the care they received. These studies focussed on people with an existing diagnosis of a mental illness or mental disorder accessing emergency care for acute MSK pain presentations. The studies investigated a variety of backgrounds including culturally and linguistically diverse, physiological and mental wellbeing, socioeconomic impact and factors such as re-attendance rates.

### Literature Synthesis

After data extraction, and synthesis of findings, we identified four main themes within the study outcomes which we will further expand upon. These included: the prevalence of concurrent pain and mental health presentations to ED; the screening and triage process for people with MKS pain when presented to ED settings; the impact of mental illness on diagnostic overshadowing and emergency staff attitude. The summary of studies is provided in the supplementary tables. The existing literature in the field focuses on a variety of MSK presentations from traumatic injuries to exacerbation of pre-existing MSK concerns and the various assessments and screening tools used to provide pain relief via medication and non-pharmacological approaches including follow up care in the community that affect continuity of care and improve patient outcomes post ED presentations.

#### 1. Concurrent pain and mental health disorder presentations to ED

Acute pain management, which accounts for nearly 80% of ED visits [18], remains a critical issue and often presents as intertwined with mental health issues and other physical chronic conditions. This theme emerged as an overview of the available literature addressing pain related challenges within ED settings, such as, overuse, psychiatric comorbidities, and the ongoing need for effective management strategies. Studies reporting the overuse of ED services by patients with chronic conditions such as lower back pain and recurrent abdominal pain, underscoring the necessity for person centred care management approaches to redirect these cases to more appropriate care settings [18-20].Concurrently, psychiatric comorbidities such as anxiety, depression, and PTSD significantly impact pain experiences and treatment outcomes in ED settings, emphasising the crucial role of integrating mental health considerations into pain management protocols [21-23].

Moreover, disparities in pain management experiences among different demographic groups underscore the importance of targeted interventions to promote equitable healthcare practices and address systemic inequities [15, 24, 25]. Existing literature links multiple emergency presentations in the context of MSK pain and the likelihood of experiencing a concurrent mental illness [22; 24; 26; 15; 20], particularly an existing post-traumatic stress disorder (PTSD) diagnosis and chronic pain, living in the inner city and low income [21] experiencing increasing vulnerabilities due to low income and lack of social support.

#### 2. Screening and Triage of Patients with MSK Pain and Mental Health Disorders in ED

The evidence highlights that pain assessment and management, MSK disorders, and psychiatric presentations are concurrent challenges commonly encountered by ED and triage clinicians [27]. For acute pain presentations at ED, the triage staff have to balance pain management, while prescribing pain-reducing medications (e.g., opioids) with potential risks associated, including dependency [28]. The role of triage staff making care decisions is pivotal in the management of acute pain presentations at the ED and plays an important role in preventing further deterioration and mitigating associated care burden.

The experiences of people with a pre-existing mental health condition attending ED have revealed care pathway disparities that can lead to future presentations, repeated attendances, exploration and delays in appropriate intervention [29]. Discrimination and bias by triage staff may contribute to poorer outcomes and inequitable treatment and care pathways [30, 31]. Further, the influence of psychosocial factors on recovery of acute MSK presentations underlines an urgent need for comprehensive assessments and suitable interventions tailored to the person and their circumstances [16].

#### 3. Diagnostic Overshadowing of Patients with MSK and mental health disorders in ED setting

The term, ‘diagnostic overshadowing” is used to detail the clinical undertakings of health professionals when assuming that the behaviour of a person with cognitive disabilities is the primary driver behind their presentation of disability, without exploring/identifying other biological or social determinant drivers [32]. The practice of diagnostic overshadowing spans all care settings, but the practice of, and consequences from, such practice could, probably does, lead to a lack of consideration of many common comorbid psychopathologies in persons seeking care for physical ailments and thus, the delivery of inappropriate care with increased risk to the person…and undue burden to the treating workforce.

Across included and reviewed literature, diagnostic overshadowing was identified and linked to discrimination and bias displayed by triage staff [54, 56, 59, 63, 64]. The synthesis of the studies demonstrated the impact on patients living with mental health concerns, their pain management and care pathways [33 - 35], clinical decision making, [36 - 38]. This led to poorer outcomes for people attending along with inequity to treatment pathways and increased chances of representation which is indeed commonly seen in clinical practice.

An existing review of the literature looking at ED presentations relating to pain issues whilst living with a mental illness or disorder, suggests that people with mental health issues typically receive a different standard of care, leading to increased rates of undiagnosed conditions, poorer outcomes, which in turn prevents them for accessing further intervention, increasing the likelihood of premature mortality [32] [39]. Addressing diagnostic overshadowing within triage settings requires acknowledgement, trauma - informed care, self-reflection and collaboration between physical and mental health multidisciplinary teams [40, 32, 34, 36, 37, 38]. While healthcare professionals do not intentionally discriminate against people with a background of mental health issues, it is important to build trust and rapport and apply the same diagnostic rigor they would apply to other people without a background of MH issues.

#### 4. Emergency Staff Attitude Towards Patients with Mental Health Disorders

This theme emerged from studies identifying the mental health stigma associated with attending an ED and the challenges faced by ED clinicians to understand the attitudes, perceptions and experiences of triage clinicians offering insight into healthcare delivery. There are diverse approaches used by different ED clinicians to address pain management challenges within ED environments to address acute and chronic pain effectively [41]. Across all these diverse approaches, healthcare providers’ attitudes and treatment recommendations are impacted by serious mental illness or having disadvantaged backgrounds in individuals presenting to ED with acute MSK pain. Provider perceptions, biases, and conceptualisations have shown to significantly influence care delivery and patient outcomes in these populations [18, 42, 43]. These studies suggest that implicit biases lead to less effective treatment recommendations, emphasizing the critical need to address these biases to achieve equitable treatment outcomes for all patients, especially those from disadvantaged backgrounds.

The literature highlights the pervasive stigma towards individuals with mental health conditions within ED settings. Stigma impacts patient experiences and access to appropriate care pathways, necessitating interventions to promote inclusivity and reduce discrimination [43]. The concerns identified in the literature include prolonged stays in the emergency department (ED) for individuals with mental health conditions, often attributed to extended assessments or systemic issues [43]. Patients and families frequently report feeling they are given lower priority during triage when seeking mental health services in the ED [33]. Furthermore, individuals with mental health conditions often believe their physical complaints are not treated as seriously as those of patients without a mental health history [35]. Additionally, there is a perception that individuals with mental health conditions may be seen as manipulative by the treating team, which further impacts their experience in the ED [44].

Training and preparedness among emergency nurses in generalist ED settings influence interactions with patients presenting with psychiatric comorbidities. This underscores the importance of tailored training programs to equip ED staff with the necessary skills and sensitivity [45]. Efforts to involve patients in their treatment decisions and improve clinician training are identified as crucial strategies to mitigate stigma and bias in emergency healthcare settings [30, 31, 33, 35].

In summary, these collectively highlight the complexity of pain management and mental health care within ED settings, placing an emphasis on the need for comprehensive approaches to address clinician perspectives, training gaps, stigma reduction, and tailored care pathways.

## Discussion

This non-systematic literature review highlights that individuals living with mental health illness have different experiences of accessing emergency care for acute MSK pain within EDs than those not living with mental health illness, potentially impacting their care pathways after an acute MSK pain.

Individuals with acute MSK pain and mental health emergencies often face stigma with the possibility that diagnostic overshadowing could influence assessment, triage, and management. These factors tend to negatively impact outcomes by influencing access to the most appropriate care pathways, due to clinician biases, both conscious and unconscious alongside other modifiable factors (e.g. gaps in training and a lack of multidisciplinary input).

Effective management requires a wholistic, time-efficient, approach integrating pain relief with mental health support, whilst considering social factors, and carefully balancing medications to avoid worsening these conditions [46, 47]. Recognising these complex needs and ensuring comprehensive assessments and care plans are imperative to address the physical symptoms of, and mental health challenges experienced by, many patients.

It is noted that the difficulties with accessing treatment for physical symptoms tend to be further impacted by busy triage settings with limited resourcing and time constraints as the presentations can be challenging to differentiate clinically. But, collaborative care models, featuring multidisciplinary teams comprising ED physicians, nurses, and allied health practitioners, pain specialists, mental health liaisons, and primary care providers, have shown promise in ensuring comprehensive care [18, 26, 28, 38].

The variations across perceptions of wellness and illness have been documented and demonstrate strong links between emotional regulation, existing mental health diagnoses or underlying vulnerabilities and pain, and how these influence recovery. There are frameworks to highlight the complexities that different people present with in case of a health crisis (physical or mental health), which provide foundation for clinicians to better understand the many causes of and contributors to illness (e.g. individual traits, genetics, social influences, variances in cultural perceptions of disease and views on recovery). Understanding the interplay of these factors informs care decisions when made in consultation with the person seeking/requiring support and considering their previous, current, and future psycho-social circumstances.

## CONCLUSION

Concurrent presentations of pain and mental health disorders in the ED highlight the need for a nuanced understanding of these complex interactions. By focusing on integrated assessment, collaborative care models, education, community resources, and patient-centred approaches, healthcare providers can better support these vulnerable patients.

Addressing these challenges holistically not only improves immediate care but also promotes long-term health and well-being… on a patient-by-patient basis.

## Data Availability

Narrative review - table attached in main manuscript.

